# Double blind placebo-controlled trial for the prevention of ulcerative colitis relapses by β-fructan prebiotics: efficacy and metabolomic analysis

**DOI:** 10.1101/2022.01.16.22269376

**Authors:** Rosica Valcheva, Heather Armstrong, Ognjen Kovic, Michael Bording-Jorgensen, Simona Veniamin, María Elisa Pérez-Muñoz, Natasha Haskey, Melissa Silva, Farhad Peerani, Karen Wong, Dina H. Kao, Sander Veldhuyzen Van Zanten, Karen I. Kroeker, Deanna L. Gibson, Eytan Wine, Michael Gänzle, Jens Walter, Levinus A. Dieleman

## Abstract

**Background:** Ulcerative colitis (UC) is associated with altered intestinal microbiome (‘dysbiosis’), most significantly, reduced strict butyrate-producing anaerobes and increased facultative anaerobes. Inulin-type prebiotics appear to reduce and prevent colitis in preclinical studies and small clinical trials. However, these results need to be validated in randomized controlled clinical trial (RCT) studies.

**Aims:** The aim of this RCT study was to assess the efficacy of β-fructans (oligofructose and inulin) in preventing relapses in UC patients in clinical remission as well as identify potential mechanisms of activity.

**Methods:** Adult UC patients in clinical remission (total Mayo score ≤ 2) were randomized to supplement their diet with 15g/d of either β-fructans (oligofructose and inulin; Synergy1/Prebiotin) or placebo (maltodextrin) for 6 months. Partial Mayo scores, medications, adverse events and intervention compliance were monitored monthly. Fecal specimens were collected throughout the study to measure fecal calprotectin (FCP), along with stool metabolites.

**Results:** Eighty-nine UC patients in clinical remission were randomized to β-fructans (n=43) or placebo (n=46). Of those, 11 participants never started, 2 were withdrawn, and 76 were included in the study population (β-fructans n=35; placebo n=41). Although there was no difference in symptomatic clinical flare (β-fructans n=11 flare; placebo n=10 flare, P=0.60), patients randomized to oligofructose and inulin showed only a 3-fold median FCP increase versus 17-fold in the placebo group (P=0.038). Interestingly, antibiotics and serotonin reuptake inhibitors significantly increased the relative risk (RR) of flare (RR 3.321, 95% Cl 2.005 to 5.344, P < 0.0001). β-Fructan intake significantly increased anti-inflammatory fecal metabolites (arabinose, L-arabitol, 5-oxo-D-proline).

**Conclusions:** Although oligofructose and inulin did not prevent symptomatic relapses in UC patients, their oral administration significantly reduced the severity of biochemical relapse compared to placebo which was further associated with increased anti-inflammatory metabolites.

## Introduction

Inflammatory bowel diseases (IBD), including Crohn disease (CD) and ulcerative colitis (UC), are common and severely debilitating chronic intestinal disorders affecting 1 in 150 people in developed nations and incidence is rising globally.^1-4^ The etiology of IBD remains poorly understood; however, associated risk factors include genetic susceptibility, urban lifestyle, dietary factors, heightened hygiene, and gut microbes.^3, 5-10^ Dysbiosis, or altered microbiome, is associated with IBD^11-13^ and can be impacted by diet.^14-17^ In particular, non-digestible carbohydrates (fiber and resistant starch) pass through the small intestine undigested; instead, some are fermented by gut microbes in the large bowel, representing a mechanistic link between diet and microbes.^18-21^ Fiber fermentation by microbes provides intestinal host cells with carbon and energy. Fermentation of dietary fibers produces gas, lactate, and short chain fatty acids (SCFAs)^21^ such as butyrate, acetate, and propionate in the colon.^22, 23^ SCFAs are absorbed and have beneficial physiological effects on shaping the gut microenvironment,^23^ immune response,^24^ and energy metabolism.^25-29^

The non-digestible inulin-type β-fructans are classified as prebiotics as they have been shown to beneficially alter activity of gut microbiota.^30^ Their clinical potential for use in IBD has been supported through studies demonstrating their ability to reduce intestinal inflammation in rodent colitis models,^31^ induce clinical response in mild UC patients associated with increased butyrate production,^32^ reduced fecal calprotectin (FCP) in UC patients,^33^ and decrease pro-inflammatory cytokines TNF-α and IL-1α mRNA expression in their colonic mucosa.^34^ Controversial results are reported in mild-moderate CD where inulin-type β-fructans were associated with induction of anti-inflammatory cytokines in CD mucosa, but with no reported clinical benefit.^35^ The efficacy of prebiotics in managing inflammation in IBD however, is not as well documented and it remains disputed whether their activity relates to the stimulation of specific members of the intestinal microbiota, or to metabolic alterations such as an increased production of SCFAs.^30, 36^ A reoccurring issue with such studies is that the duration of treatment may be too short to assess clinical and endoscopic improvement, and often, the colonic microbiota were not analyzed.

The present randomized controlled trial (RCT) study aimed to examine safety, tolerability and efficacy, and to define mechanisms underlying the activity of inulin-type β-fructans for use in prevention of UC relapse in patients in clinical remission and who had a documented relapse in the preceding 18 months. Remission adult UC patients were randomized to supplement their diet with 15 g/d of either prebiotics (β-fructans) or placebo (maltodextrin) for 6 months. Fecal and serum specimens were collected throughout the study and outcome measurements included clinical parameters, FCP, and metabolites, which were associated with disease activity as defined by FCP change from baseline to month 6, or at relapse. Consumption of inulin-type β-fructans did not prevent clinical relapse in UC patients who were in clinical remission, however it significantly reduced risk of relapse, defined by increased FCP (subclinical or biochemical colonic inflammation).

## Methods

### Study Settings and Design

This study was a single-center, double-blind, randomized, parallel, placebo controlled clinical trial (RCT) comparing the effectiveness of β-fructans (Synergy-1/Prebiotin; combination of oligofructose and inulin) versus maltodextrin (as placebo) to prevent clinical relapse among patients with inactive UC. The study, conducted at University of Alberta (Edmonton, Canada), occurred between 4 July 2016 and 1 December 2020. All research procedures performed in this trial were in strict accordance with a predefined protocol and adherence to international GCP guidelines and Declaration of Helsinki. The research protocol was approved by Health Research Ethics Board (Study ID Pro00041938) at the University of Alberta and Natural Health Directorate at Health Canada. The study is publicly accessible at the U.S. National Institute of Health database (clinicaltrials.gov identification number NCT02865707).

### Patients, inclusion and exclusion criteria

Patients aged 18-65 years, with clinical and endoscopic confirmed remission UC, Mayo Clinical Score ≤2 on the 12-point Mayo scale at baseline,^37^ endoscopic score 0 or 1, and with no documented clinical relapse within 18 months before the study, were eligible to participate. Inclusion criteria were treatment on stable doses of 5-aminosalicylic acid (5-ASA) for at least 2 weeks, azathioprine and/or biologics (infliximab, vedolizumab) for at least 2 months prior to screening, or no medication for at least 1 week prior to the start of the trial, and negative tests for Clostridoides difficile toxin, stool pathogens, and pregnancy.

Exclusion criteria included use of oral or steroids 4 weeks prior to the screening, topical 5-ASA and steroids within 1 week, use of methotrexate or 6-mercaptopurine, use of antibiotics within 2 months, and use of anti-diarrheal agents within 3 days of the screening visit. Patients with significant chronic disorders (severe cardiac disease, renal failure, severe pulmonary disease or severe psychiatric disorder), any active infection, or pregnancy were also excluded.

Patients matching the inclusion and exclusion criteria were approached and only those who provided written informed consent were recruited into the study.

### Study Intervention and Procedures

All study products were supplied by BENEO GmbH (Germany) in sachets. The study intervention consisted of two arms: 1) β-fructans (Prebiotin/Synergy1^®^, oligofructose and inulin in ratio 1:1 manufactured by BENEO GmbH, Germany, and kindly provided by Jackson GI Medical LLC, Harrisburg, PA, USA) 15 g/day; and 2) Placebo (Agenamalt 22.222 maltodextrin DE19 manufactured by AGRANA Beteiligungs-AG, Austria) 15 g/day. Maltodextrin was chosen as placebo since it is thought to be fully metabolized in the small intestines and would have no effect on the colonic gut microbiota fermentation.^38, 39^ Participants were advised to dissolve the powder products in 200 ml warm liquid (water or other beverage) and drink it during meal. To improve tolerability, participants began with an adjustment period of 2 weeks consuming half of total daily product’s amount (7.5 g/day).

A summary of study procedures is shown in **Figure 1**. At screening, consented patient underwent a sigmoidoscopic and physical examinations to confirm eligibility and total Mayo clinical score was calculated. If patients qualified, then biopsies were collected at 20-25 cm from the anal verge. In addition, demographics [including age, sex, height and weight to calculate body mass index (BMI)], full medical history, UC disease location, list of current medications and supplements were recorded. The same procedures were performed at 6 months, or at relapse. Patients were randomized in 1:1 fashion to one of the treatment arms using random block sizes of 2 and 4, stratified based on sex (males versus females). The randomization sequence was generated by University of Alberta Epidemiology Coordinating and Research (EPICORE) Centre and embedded into REDCap (Research Electronic Data Capture)^40^ electronic case report form (eCRF) system. Participants were required to maintain a stable dietary pattern, monitored by web-based Diet History Questionnaire II (DHQII)^41^ completed at baseline and 6-months/relapse as well as to remain on the same dosage and type of medication for UC maintenance (confirmed at month 1, 2, 3, 4, 5, 6 or at relapse). During the study period participants provided stool at baseline (month 0), month 1, month 3, and month 6 or disease relapse. Blood samples for haematology and chemistry were collected at baseline, month 3 and month 6 or at relapse. Incidence and severity of all adverse events were captured during the follow-up interviews every month. The adverse events were classified by the Investigator with regards to possible relationship to treatment (unrelated, unlikely, possible or probable) and according to severity (mild, moderate, severe). Compliance to study supplementation was defined as having used ≥ 85% of the provided sachets. It was assessed at month 3 and month 6, or at relapse.

**Figure 1:**
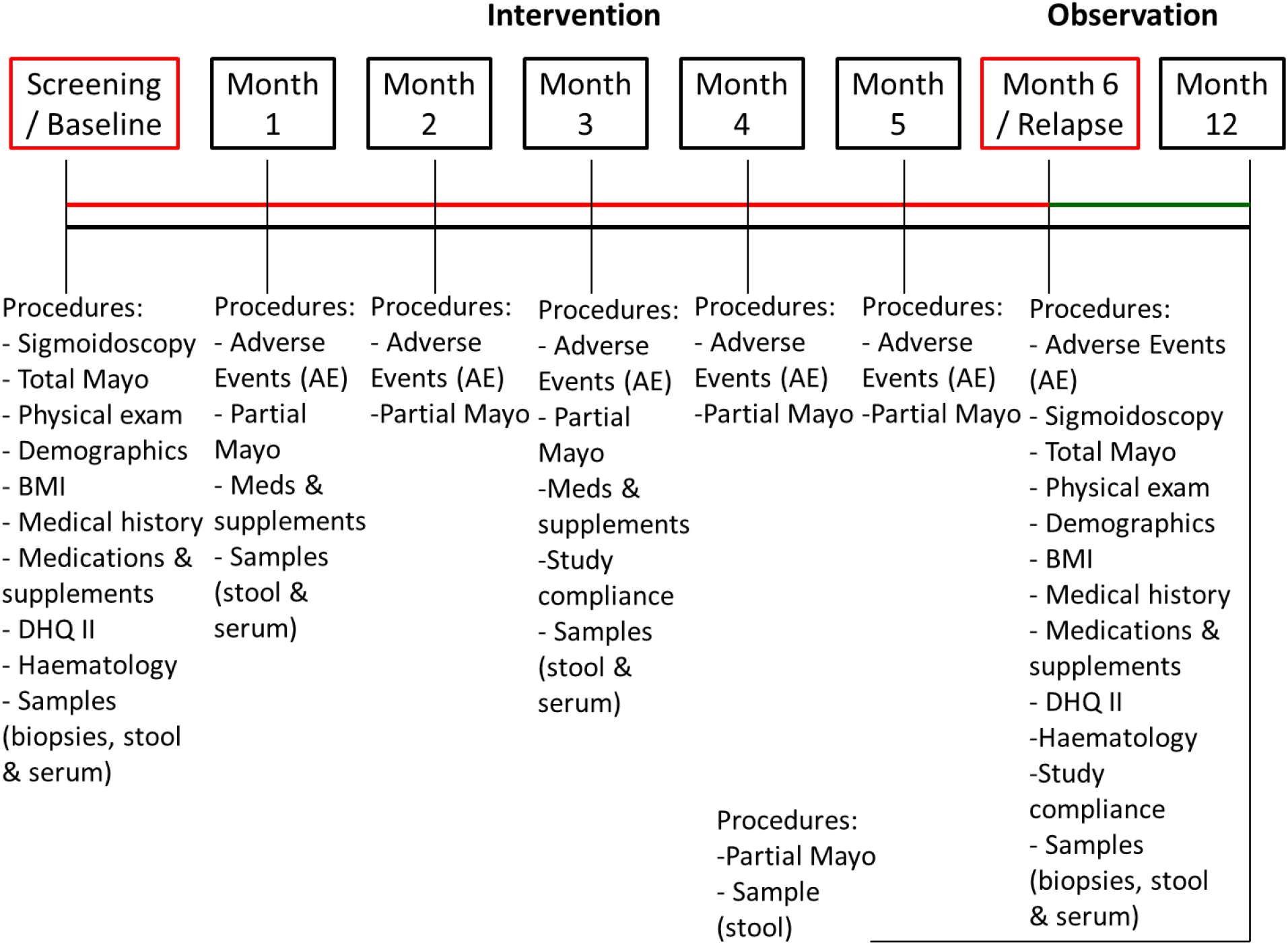
Summary of study procedures and collected samples

### Outcome Measures

The primary outcome measure was patient clinical relapse rate over 6 months of active supplemental intervention. Clinical relapse was based on partial Mayo score ≥ 3.^42^ Key secondary outcomes were 1) changes in FCP at month 6 or at relapse, versus baseline FCP; 2) changes in partial Mayo clinical score at month 6 or at relapse, versus baseline Mayo; 3) changes in endoscopic score at month 6 or at relapse, versus baseline endoscopic score; and 4) time to relapse.

Treatment failure (disease relapse) was defined as an increase in UC disease activity measured as 1) a partial Mayo score ≥ 3; or 2) FCP concentration > 250 µg g^-1^.^42^ Treatment response was defined as maintained remission based on the FCP at month 6 versus month 0 (FCP <250 µg g^-1^ and/or FCP rise of less than 100 at endpoint). For participants who withdrew before the time of treatment assessment (month 6 or relapse) or were lost to follow-up missing data were imputed using last observation carried forward.

Additional safety outcome measures included number of patients experiencing a serious adverse event (SAE) during treatment period as well as stool culture testing positive for intestinal pathogens. Safety assessment also included measures of C-reactive protein (CRP), blood haematology (hemoglobin, MCV, platelets, WBC, albumin), iron deficiency [iron, total iron binding capacity (TIBC), ferritin iron saturation index], serum liver and kidney markers (serum creatinine, aspartate aminotransferase (AST), alanine transaminase (ALT), gamma-glutamyl transferase (GGT), alkaline phosphatase (ALP), bilirubin) as well as 25-hydroxy vitamin D at baseline versus month 3 and endpoint (month 6 or relapse). Tolerability outcome measures were defined based on total treatment emergent adverse events (TEAE) versus such TEAE leading to withdrawal.

### Sample Size Determination

Data from our pilot study in UC patients with active disease supplemented with β-fructans (Synergy1^®^) to stable dose 5-ASA treatment for 9 weeks demonstrated a 77% response in 15 g/day dose versus 33% in a lower dose of 7.5 g/day.^32^ Based on this study outcome we have chosen a dose of 15 g/day to be tested in the current study with an estimated 30% difference in the incidence of UC patients experiencing clinical relapse by 6 months. A total of 84 patients, 42 in each arm, were required to detect a difference of 30% with a power of 80% using a two-sided p=0.05 level test. With an anticipated dropout rate of 20%, the overall sample size for this trial was proposed be 100 patients.

### Metabolomics

Untargeted metabolites were isolated from patient stool samples as directed by the Calgary Metabolomics Research Facility, University of Calgary. On ice, 100-200mg of stool were measured into 1.5mL Eppendorf tubes and weights were recorded. Five volumes of pre-chilled methanol was added to each sample, then homogenized at max speed in the MPBio Fastprep24 bead beater for 1 min. Samples were incubated on ice for 30min then centrifuged at max speed, 4c, for 10min. Supernatants were processed for µM concentration of metabolites by Calgary Metabolomics Research Facility staff.

### Quantification and Statistical Analysis

#### Statistical Analysis

Groups were compared using paired Wilcoxon *t*-test (two-tailed) analysis, ANOVA, or Kendall, depending on the relevant question, using GraphPad Prism. A p-value of <0.05 was considered significant in all cases and all error deviations are described by ± SEM.

## Results

### Recruitment and Participant Flow

We screened 113 potential participants, of whom 34 failed to meet the eligibility criteria. **Figure 2** outlines the patient flow in the recruitment, intervention allocation and reasons for exclusion. Of the 89 patients who successfully passed screening, 43 were randomly assigned to the β-fructans arm and 46 to the placebo arm. One participant developed allergic reaction to β-fructans, manifested by skin rash, shortly after starting the intervention (24-48h), and was immediately withdrawn. Two participants, one in each group, had to be withdrawn from the study as they became ineligible (one had recurrent *C. difficile* infection, and one had protocol violation related to medication and failure to randomization). Finally, 10 participants never started the study procedures and were lost to follow-up. Thus, the safety study population included 76 subjects with 35 in the β-fructans arm and 41 participants in the placebo arm who participated in at least one follow-up study visits. The demographics and baseline characteristics of the safety population are shown in **Table 1** and were similar across the treatment groups. Overall, 57% of the participants were women and mean age at entry was 45.0 ± 13.4 years. The median body mass index (BMI, kg m^-2^) was 27.0 (IQR 24.5-30.0) suggesting a population mostly in the overweight category. Baseline median FCP concentrations were 113.8 µg g^-1^ (IQR 48.9 - 471.1) with about one third of the participants in each arm having elevated concentrations > 250 µg g^-1^ (β-fructans n=9, 26%; placebo n=13, 31%) though exhibiting no clinical symptoms. A history of pan-colitis accounted for 60% of all UC. At baseline 33% of the participants were taking biologic medications, 41% were only on 5-ASA and 16% were on no medication related to UC.

**Table 1.** Baseline demographics of the Safety study population

| Characteristic | β-Fructans<br>(N = 35) | Placebo<br>(N = 41) | Total<br>(n = 76) |
| --- | --- | --- | --- |
| Sex (Female) n (%) | 19 (54%) | 24 (58%) | 43 (57%) |
| Mean age at entry (yr) ± SD; range | 44.1 ± 13.7<br>(20 - 65) | 45.2 ± 13.8<br>(21 - 68) | 45.0 ± 13.4<br>(21 - 68) |
| BMI (kg/m<sup>2</sup>) median (IQR) | 26.7<br>(23.1 - 29.7) | 27.3<br>(24.9 - 30.5) | 27.0<br>(24.5 - 30.0) |
| Disease Location |  |  |  |
| Left-sided colitis, n, (%) | 11 (31%) | 13 (32%) | 24 (32%) |
| Pancolitis, n, (%) | 23 (66%) | 23 (56%) | 46 (60%) |
| Proctitis, n, (%) | 1 (3%) | 5 (12%) | 6 (8%) |
| Medications |  |  |  |
| No Medication, n, (%) | 7 (20%) | 5 (12%) | 12 (16%) |
| 5-aminosalicylic acid (5-ASA) only, n, (%) | 14 (40%) | 17 (42%) | 31 (41%) |
| Immunosuppressants (Imuran), n, (%) | 1 (3%) | 1 (2%) | 2 (2%) |
| Biologics, n, (%) | 4 (11%) | 6 (15%) | 10 (13%) |
| Combined 5-ASA/Imuran, n, (%) | 1 (3%) | 5 (12%) | 6 (8%) |
| Combined 5-ASA/Biologics, n, (%) | 5 (14%) | 4 (10%) | 9 (12%) |
| Combined 5-ASA/Imuran/Biologics, n, (%) | 2 (6%) | 1 (2%) | 3 (4%) |
| Combined Imuran/Biologics, n, (%) | 1 (3%) | 2 (5%) | 3 (4%) |
| Fecal calprotectin (μg/g) median (IQR) | 113.3<br>(48.7 - 333.9) | 129.1<br>(47.8 - 539.5) | 113.8<br>(48.9 - 471.1) |
| C-reactive protein (l) median (IQR) | 1.4 (0.5 - 2.6) | 2.3 (0.5 - 7.0) | 1.5 (0.5-5.3) |
| Comorbidities |  |  |  |
| No comorbidities, n, (%) | 11 (31%) | 13 (32%) | 24 (32%) |
| Asthma, n, (%) | 6 (17%) | 3 (7%) | 9 (12%) |
| Celiac disease, n, (%) | 3 (9%) | 0 (0%) | 3 (4%) |
| Chronic obstructive pulmonary disease, n, (%) | 0 (0%) | 3 (7%) | 3 (4%) |
| Depression / Anxiety, n, (%) | 5 (14%) | 4 (10%) | 9 (12%) |
| Diabetes Mellitus Type 2, n, (%) | 3 (9%) | 1 (2%) | 4 (5%) |
| Gastro-esophageal reflux disease, n, (%) | 1 (3%) | 7 (17%) | 8 (11%) |
| Hypercholesterolemia, n, (%) | 0 (0%) | 5 (12%) | 5 (7%) |
| Hypertension, n, (%) | 3 (9%) | 8 (20%) | 11 (14%) |
| Hypothyroidism, n, (%) | 3 (9%) | 2 (5%) | 5 (7%) |
| Multiple sclerosis, n, (%) | 1 (3%) | 0 (0%) | 1 (1%) |
| Pancreatitis, n, (%) | 1 (3%) | 1 (2%) | 2 (3%) |
| Primary schlerosis cholangitis, n, (%) | 2 (6%) | 1 (2%) | 3 (4%) |
| Other, n, (%) | 4 (11%) | 4 (10%) | 8 (11%) |

**Figure 2:**
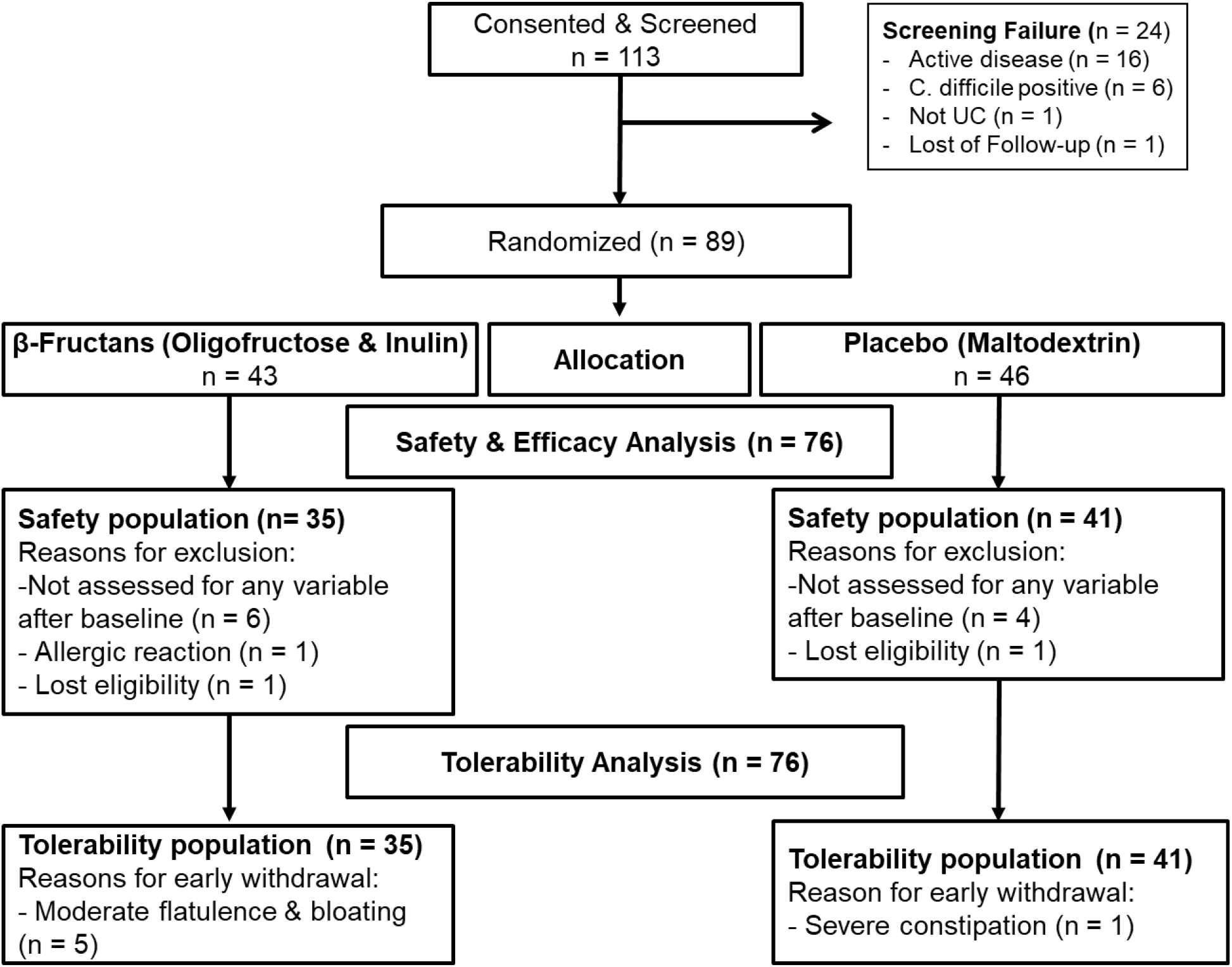
Patient flow in the recruitment, intervention allocation and reason for exclusion. Of the 89 patients who successfully passed screening, 43 were randomly assigned to receive β-fructans intervention and 46 to the placebo arm. 13 patients either never started the trial or were lost to follow-up therefore, the intention-to-treat study population included 76 subjects with 35 in the β-fructans arm and 41 in the placebo arm. Further patients withdrew their consent during the study, therefore in total 65 participants completed the study including 29 in the β-fructans arm and 36 in the placebo arm.

### Adverse Events and Safety Parameters

All symptoms occurring during the study were documented and rated for severity and potential relationship with the study products. **Table 2** summarizes all treatment emergent adverse events (TEAE) described by the study population during the course of 6 months intervention. Significantly more participants in the β-fructans group reported the onset of new symptoms (n=25, 71%, p=0.006) in comparison to such TEAE in the placebo population (n=16, 39%). Inulin type β-fructans intake was associated with mild or moderate flatulence and bloating reported by 51% of the study population (n=18) and thus was considered as a treatment related adverse event. Five participants elected withdrawal due to this TEAE. FCP analysis determined that this TEAE was not associated with increased subclinical inflammation [baseline FCP median 76.1 µg g^-1^ (IQR 45.4 - 589.3) versus early withdrawal FCP median 42.9 µg g^-1^ (IQR 21.8 - 92.3), p = 0.625] (**Supplementary Figure 1**). For the remaining patients experiencing flatulence and bloating, this symptom resolved within 2-8 weeks. Other gastrointestinal symptoms, such as pyrosis, diarrhea or constipation were insignificant and equally reported by both treatment groups, although resulted in one participant withdrawal in the placebo group. Besides gastrointestinal-associated TEAEs, 12 patients, 6 in each treatment group, reported acute infections (respiratory, tooth abscess, STI) and 9 were treated with antibiotics (β-fructans n = 4; placebo n = 5). Finally, most haematologic and serum parameters remained in the normal ranges and did not significantly change across groups from baseline (**Table 3**). CRP levels remained low and BMI index did not alter after 6 months prebiotic intervention.

**Table 2.** Summary ofTreatment Emergent Adverse Events Recorded in the Safety Population

| Characteristic | β-Fructans<br>N = 35 | Placebo<br>N = 41 | n p-value |
| --- | --- | --- | --- |
| TEAEs, n, (%), E | 25 (71%) 33 | 16 (39%) 26 | 0.006 |
| Anxiety/depression, n, (%), E | 1 (3%) 1 | 1 (2%) 1 | >0.999 |
| Constipation, n, (%), E | 0 (0.00) 0 | 1 (2%) 1 | >0.999 |
| Diarrhea, n, (%), E | 0 (0.00) 0 | 2 (5%) 2 | 0.496 |
| Dry mouth, n, (%), E | 0 (0.00) 0 | 1 (2%) 1 | >0.999 |
| Eczema, n, (%), E | 0 (0.00) 0 | 1 (2%) 1 | >0.999 |
| Flatulence/bloating mild, n, (%), E | 11 (31%) 11 | 4 (10%) 4 | 0.022 |
| Flatulence/bloating, moderate, n, (%), E | 7 (20%) 7 | 0 (0.00) 0 | 0.003 |
| Gallbladder stones, n, (%), E | 0 (0.00) 0 | 1 (2%) 1 | >0.999 |
| Headache, n, (%), E | 1 (3%) 1 | 0 (0.00) 0 | >0.999 |
| Pyrosis, n, (%), E | 2 (6%) 2 | 2 (5%) 2 | >0.999 |
| Hemorrhoids, n, (%), E | 0 (0.00) | 1 (2%) 1 | >0.999 |
| High blood pressure, n, (%), E | 1 (3%) 1 | 0 (0.00) 0 | >0.999 |
| Infection (bacterial, viral), n, (%), E | 6 (17%) 8 | 6 (15%) 7 | >0.999 |
| Insomnia, n, (%), E | 0 (0.00) 0 | 1 (2%) 1 | >0.999 |
| Joint pain, n, (%), E | 0 (0.00) 0 | 1 (2%) 1 | >0.999 |
| Seizure, n, (%), E | 1 (3%) 1 | 0 (0.00) 0 | >0.999 |
| Surgery for Carpal Tunnel Syndrome, n, (%), E | 1 (3%) 1 | 0 (0.00) 0 | >0.999 |
| Weight gain, n, (%), E | 0 (0.00) 0 | 2 (5%) 2 | 0.496 |
| Weight loss, n, (%), E | 0 (0.00) 0 | 1 (2%) 1 | >0.999 |
| Serious TEAEs, n, (%), E | 0 (0.00) 0 | 0 (0.00) 0 | >0.999 |
| Severe TEAEs, n, (%), E | 0 (0.00) 0 | 0 (0.00) 0 | >0.999 |
| Related TEAEs, n, (%), E | 18 (51%) 18 | 0 (0.00) 0 | <0.0001 |
| TEAEs Leading to Withdrawal, n, (%), E | 5 (14%) 5 | 1 (2%) 1 | 0.089 |
| TEAEs Leading to Death, n, (%), E | 0 (0.00) 0 | 0 (0.00) 0 | >0.999 |

E: Number of Events; n: Number of subjects with characteristic; TEAE: Treatment Emergent Adverse Event; %: calculated as the proportion of subjects with a characteristic versus the total number of subjects in the group ( β-fructans or placebo) safety population ( $n/N \times 100$ )

**Table 3.** Changes in blood haematology, serum liver and kidney markers in UC patients in clinical remission consuming inulin-type β-fructans for 6 months.

| Characteristic | Baseline | Endpoint | Within group P-value | Baseline | Endpoint | Within group P-value |
| --- | --- | --- | --- | --- | --- | --- |
| **Blood haematology** |  |  |  |  |  |  |
| Hemoglobin, median g L<sup>-1</sup> (IQR) | 142.0 (131 - 149) | 146.0 (133 - 154) | 0.507 | 146.0 (136 - 153) | 143.0 (137 - 155) | 0.743 |
| MCV, median fL (IQR) | 89.0 (85 - 93) | 90.0 (86 - 94) | 0.170 | 89.0 (86 - 93) | 90.0 (87 - 94) | 0.039 |
| Platelets, median 10<sup>9</sup> L<sup>-1</sup> (IQR) | 243 (211 - 294) | 250 (223 - 291) | 0.507 | 280 (253 - 318) | 289 (232 - 312) | 0.844 |
| WBC, median 10<sup>9</sup> L<sup>-1</sup> (IQR) | 5.3 (4.4 - 7.2) | 5.9 (4.7 - 7.5) | 0.087 | 6.1 (5.6 - 7.6) | 6.3 (5.1 - 8.1) | 0.478 |
| **Iron deficiency** |  |  |  |  |  |  |
| Iron, median μmol L<sup>-1</sup> (IQR) | 17.0 (11 - 21.8) | 17.5 (14 - 23.8) | 0.198 | 14.0 (10 - 21) | 17.0 (12 - 20) | 0.428 |
| TIBC, median μmol L<sup>-1</sup> (IQR) | 61.5 (55.5 - 65.8) | 59.5 (56.0 - 67.8) | 0.923 | 61.0 (54.0 - 66.0) | 58.0 (54.0 - 63.0) | 0.014 |
| **Liver and kidney function markers** |  |  |  |  |  |  |
| Albumin, median g L<sup>-1</sup> (IQR) | 44.0 (40.5 - 46.0) | 44.0 (42.0 - 46.0) | 0.241 | 43.0 (42.0 - 46.0) | 43.0 (42.0 - 46.0) | 0.669 |
| ALP, median IU L<sup>-1</sup> (IQR) | 62.0 (42.0 - 71.5) | 62.0 (46.3 - 76.8) | 0.764 | 67.0 (55.0 - 103) | 64.0 (53.0 - 84.0) | 0.385 |
| AST, median IU L<sup>-1</sup> (IQR) | 22.0 (19.0 - 26.0) | 23.0 (18.0 - 27.0) | 0.395 | 24.0 (20.5 - 29.0) | 23.0 (18.5 - 31.5) | 0.745 |
| ALT, median IU L<sup>-1</sup> (IQR) | 20.5 (16.3 - 27.8) | 24.0 (18.0 - 30.3) | 0.158 | 25.0 (16.3 - 31.0) | 20.5 (15.3 - 33.0) | 0.809 |
| GGT, median IU L<sup>-1</sup> (IQR) | 9.0 (5.0 - 11.8) | 7.5 (5.0 - 24.8) | 0.235 | 12.0 (5.0 - 32.0) | 8.0 (5.0 - 30.0) | 0.389 |
| Serum creatinine, median μmol L<sup>-1</sup> (IQR) | 76.0 (66.0 - 84.0) | 73.0 (65.0 - 84.0) | 0.873 | 73.0 (63.0 - 86.0) | 73.0 (65.0 - 86.0) | 0.882 |
| Bilirubin, median μmol L<sup>-1</sup> (IQR) | 12.0 (7.8 - 15.5) | 13.5 (10.0 - 18.0) | 0.002 | 13.0 (9.0 - 17.3) | 13.0 (10.8 - 17.5) | 0.345 |
| CRP, median mg L<sup>-1</sup> (IQR) | 1.5 (0.5 - 2.6) | 1.1 (0.5 - 4.4) | 0.569 | 2.8 (0.5 - 7.1) | 1.5 (0.5 - 6.2) | 0.176 |
| BMI (kg/m<sup>2</sup>) median (IQR) | 26.7 (23.1 - 29.7) | 26.4 (23.6 - 29.4) | 0.731 | 27.3 (24.9 - 30.5) | 28.2 (24.3 - 33.5) | 0.687 |

ALP: alkaline phosphatase; ALT: alanine transaminase; AST: aspartate aminotransferase; BMI: Body mass index; CRP: C-Reactive protein; GGT: gamma-glutamyl transferase; MCV: mean corpuscular volume; TIBC: total iron-binding capacity; WBC: white blood cells

### Primary and Secondary Outcomes

The primary outcome of the study was maintenance of clinical remission during 6 months intervention. Treatment failure (relapse) was described as an increase in clinical symptoms (partial Mayo score ≥ 3; symptomatic relapse) and/or by changes in the biochemical marker FCP (endpoint FCP > 250 µg g^-1^ or FCP rise of more than 100 µg g^-1^ in patients with high baseline FCP; FCP relapse). Linear regression analysis showed strong correlation between symptom changes (partial Mayo score) and FCP changes (F-value = 70.39, r^2^ = 0.53, p < 0.0001). During the 6-months intervention 11 individuals in β-fructans group (31%) and 10 in the placebo group (24%) experienced symptomatic relapse (chi-square 1.23, p = 0.26; **Figure 3A**). Similarly, there was no significant difference in the proportion of patients who showed biochemical relapse (β-fructans n = 10, 29%; placebo n = 16, 39%; chi-square 0.094, p = 0.76). In addition, 4 patients (β-fructans n = 2; placebo n = 2) experienced symptomatic relapse during the 6-months post-intervention observational period. Log-rank test comparing the relapse events over the 12 months period (6 months intervention followed by 6 months observation) showed no significant difference in the time to relapse between β-fructans and placebo (p = 0.92; **Figure 3B**). Based on this outcome we concluded that β-fructans intake was ineffective in improving the clinical relapse rates or time to clinical relapse. Consequently, the trial was prematurely terminated due to lack of efficacy and further recruitment was halted.

**Figure 3:**
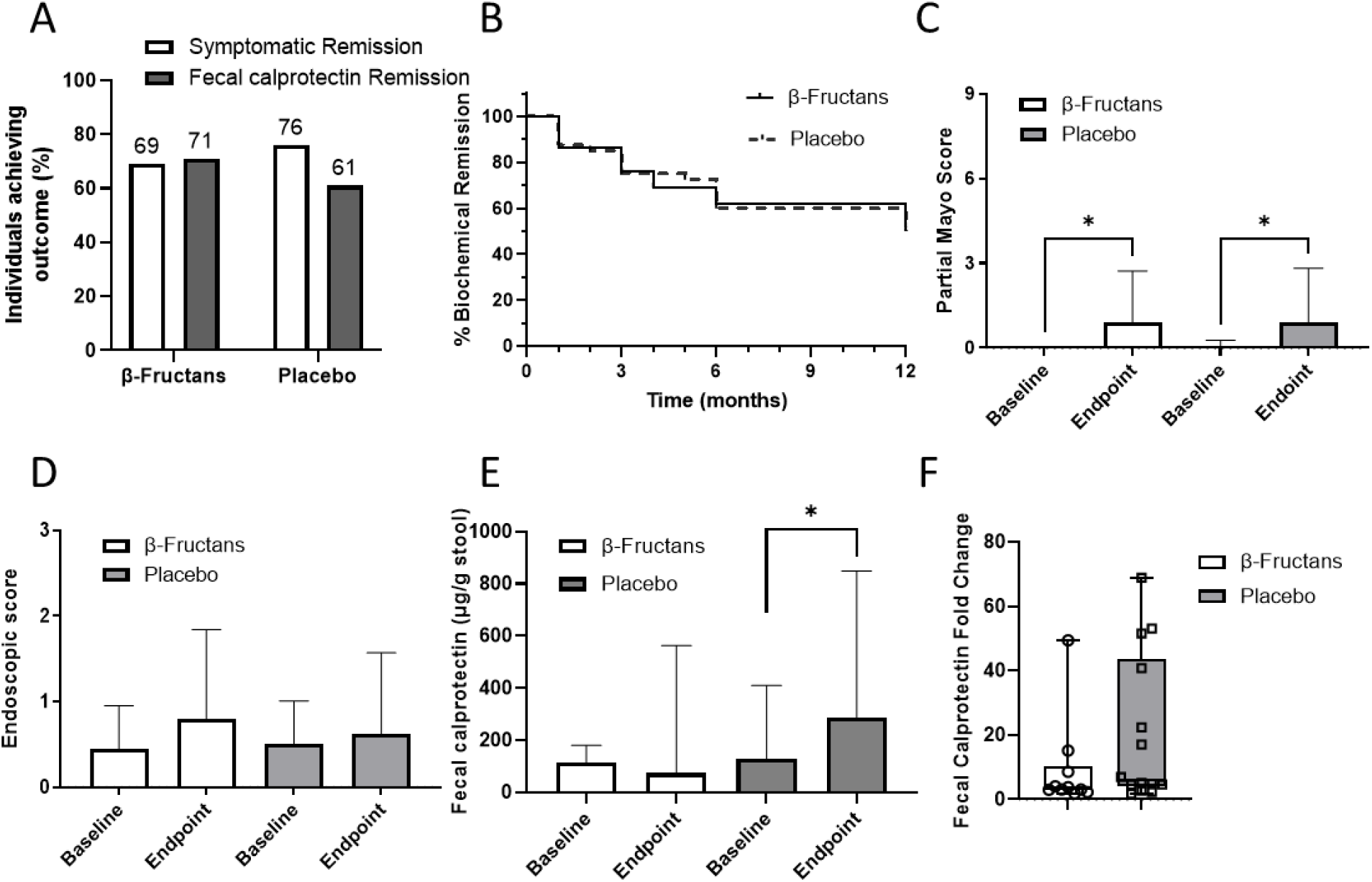
Clinical outcomes as measured in participants at baseline and endpoint.

We then assessed the secondary outcome measures, notably changes in clinical parameters partial Mayo score, colitis endoscopic score, and FCP. At baseline both treatment groups had similar partial Mayo score (β-fructans 0.0±0.0; placebo 0.0±0.2, p > 0.99), endoscopic scores (β-fructans 0.5±0.5; placebo 0.5±0.5, p > 0.99), and FCP (β-fructans median 113.3, IQR 48.7-333.9; placebo median 129.1, IQR 47.8-539.5, p = 0.63). Two-way ANOVA test showed that both β-fructans and placebo groups were characterized with a significant increase in the UC-related symptoms (partial Mayo score) versus the baseline measures (p=0.002; endpoint β-fructans 0.9±1.8; placebo 0.9±1.9; **Figure 3C**) and insignificant increase in the endoscopic scores (endpoint β-fructans 0.8±1.0; placebo 0.7±1.0; **Figure 3D**), however there was no difference between the two treatments (p>0.99). In contrast, assessment of FCP changes showed a slight but not significant reduction in FCP levels in β-fructans group (endpoint β-fructans median 73.3, IQR 22.4-1072.0, p=0.35). Placebo group demonstrated a significant increase in the FCP levels at treatment endpoint (endpoint placebo median 285.5, IQR 49.1-1122.0, p = 0.036; **Figure 3E**). Finally, there was no statistical difference in FCP absolute levels (p = 0.19), nor in FCP fold change (p = 0.10), between groups at the intervention endpoint. However, data showed that while those clinically relapsing patients randomized to β-fructans had a 3-fold median increase in their FCP from baseline (IQR 2.3-8.6), relapsing patients randomized to the placebo had a 17-fold median increase (IQR 4.6-47.0; p=0.038; **Figure 3F**); a significant difference between groups.

### Consumption of β-fructans induced significant changes in metabolites in participant stool

To understand difference between patients in relapse vs remission, we examined metabolite abundance changes over time via untargeted metabolomics of stool collected from β-fructans remission (n=15), β-fructans relapse (n=10), placebo remission (n=15), and placebo relapse (n=15) participants (**Figure 4**). At month 0, participants randomized to the β-fructans arm who remained in remission during the study, had significantly higher fecal arabinose, 3-hydroxy-3-methylglutarate, 3-hydroxybutanoic acid, α-D-glucose, D-alanine, acetoacetate, ferulate, and D-gulonic acid γ-lactone, and significantly less leucine and L-glutamic acid than those in the β-fructan arm who relapsed (**Figure 4A**). In contrast, participants randomized to the placebo arm who remained in remission had significantly higher arabinose (like β-fructans arm) and N-acetyl-L-aspartic acid, compared to those in the placebo arm who relapsed.

**Figure 4:**
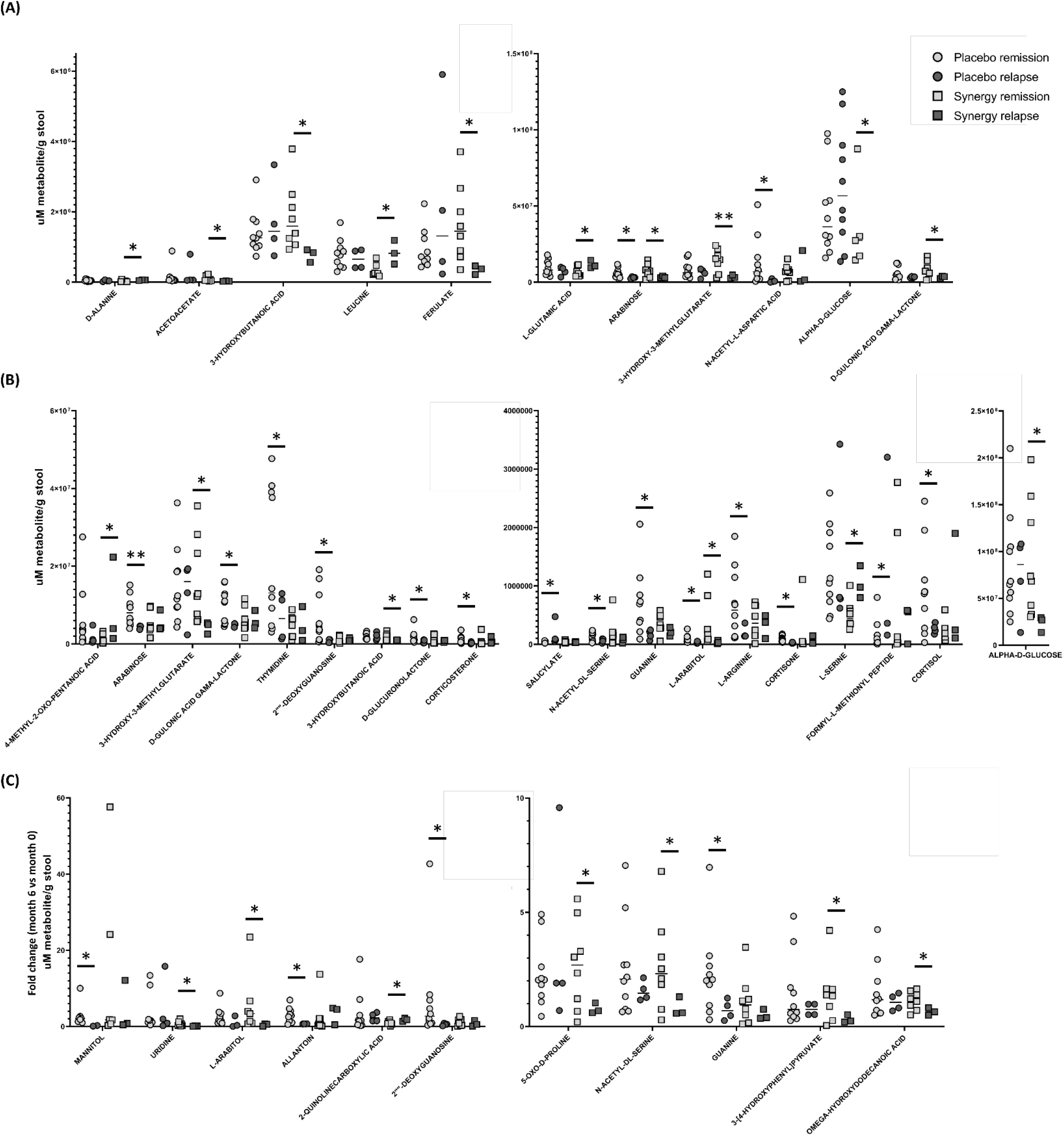
Metabolites were examined in patient stool samples isolated from synergy remission (n=8), synergy relapse (n=3), placebo remission (n=10), and placebo relapse (n=4) taken at **(A)** month 0, **(B)** month 6, and **(C)** month 6 versus month 0. **p<0.01, *p<0.05.

At month 6, participants randomized to the β-fructans arm who remained in remission had significantly higher 3-hydroxy-3-methylglutarate, 3-hydroxybutanoic acid, α-D-glucose, and L-arabitol, along with significantly less 4-methyl-2-oxo-pentanoic acid and L-serine than those in the β-fructans arm who relapsed (**Figure 4B**). In contrast, participants randomized to the placebo arm who remained in remission had significantly higher arabinose, N-acetyl-DL-serine, 2”“-deoxyguanosine, D-gulonic acid γ-lactone, thymidine, D-glucuronolactone, guanine, L-arginine, formyl-L-methionyl peptide, corticosterone, cortisone, and cortisol along with decreased salicylate compared to those in the placebo arm who relapsed.

Comparing the fold-change in metabolites from month 6 versus month 0, participants randomized to the β-fructans arm who remained in remission had significantly greater increased fold-change in N-acetyl-DL-serine, L-arabitol, 5-oxo-D-proline, 3-[4-hydroxyphenyl]pyruvate, and omega-hydroxydodecanoic acid, along with significantly decreased 2-quinolinecarboxylin acid, than those in the β-fructans arm who relapsed (**Figure 4C**). In contrast, participants randomized to the placebo arm who remained in remission had significantly higher 2”“-deoxyguanosine, mannitol, allantoin, and guanine compared to those in the placebo arm who relapsed.

## Discussion

An altered intestinal microbiome is well linked to IBD^43-45^ and mounting evidence supports the use of dietary interventions as a successful approach to re-establish microbiota in mild to moderately active IBD. Dietary fibres, in particular, are a key energy source for gut microbiota and fructan fibres have previously been shown to reduce colitis in animal models of colitis,^31, 46^ and in small clinical studies.^32-34^ Unfortunately, studies related to the clinical efficacy of fructan fibres remain limited, and those that have been published suffer from short treatment duration and small participant numbers. This RCT shows that consumption of β-fructans for 6 months by UC patients in remission was ineffective at improving clinical relapse rate or time to relapse however, β-fructans were able to reduce the overall severity of relapse by preventing subclinical inflammation or significantly reducing the amount of FCP increase during symptomatic relapse; UC patients randomized to β-fructans who relapsed had a 3-fold median increase in their FCP from baseline compared to patients randomized to placebo who relapsed (17-fold median increase; p = 0.038).

A number of metabolites were significantly altered in response to β-fructan consumption. Arabinose exerts anti-inflammatory effects in mouse models of colitis^47^ and was found to be higher at baseline in both the β-fructan and placebo arm participants who remained in remission at month 6, suggesting that high fecal arabinose may be a predictor of remission in UC patients. Unique to overall response to β-fructan consumption, those patients who remained in remission also had significantly higher L-arabitol than patients who relapsed; arabitol is produced from arabinose, although it has not previously been well defined, it can indicate overgrowth of select microbes in the gut.^48^ Increased 5-oxo-D-proline was also identified in the β-fructan remission group, compared to relapse, and has been previously shown to be significantly higher in IBD patients, linked to coping mechanisms of oxidative stress and inflammation.^49^ 3-[4-hydroxyphenyl]pyruvate, increased in those who remained in remission following consumption of β-fructans, has also previously been shown to be upregulated in IBD, although its potential role remains unknown.^50^ Lastly, hydroxydodecanoic acid, increased in those who remained in remission following consumption of β-fructans, has been shown to have anti-inflammatory effects in limited studies.^51^ Overall, the specific pathways associated with remission following consumption of β-fructans have previously been identified for their significant anti-inflammatory effects, demonstrating potential mechanistic effects that will need to be investigated in future biochemical studies.

In conclusion, this RCT study suggest that intake of 15 g/day β-fructans over a 6-month period could induce significant reduction in FCP and subclinical remission. These findings suggest that high-dose inulin-type fructan fibre supplement improves subclinical colitis demonstrating their promising potential as an adjunct treatment to aid in maintaining remission in UC. Further well-powered RCT with these prebiotics are warranted to better define personalized response to β-fructans and identify biomarkers predicting which patients are most likely to benefit from these interventions.

## Data Availability

All data produced in the present work are contained in the manuscript

## ACKNOWLEDGEMENTS

We graciously thank the patients for agreeing to participate and provide samples, as well as the IBD research coordinators, nurses, and endoscopy staff at the Zeidler Ledcor IBD Clinic, University of Alberta Hospital. We would like to acknowledge The Applied Genomic Core at the University of Alberta along with IMPACTT Microbiome and the Calgary Metabolomics Research Facility at the University of Calgary for their help in experiment preparations and technical procedures. We also thank CEGIIR for providing material support. LD was supported by CIHR and Weston Family Foundation grants and also sponsored by Jackson GI; along with Weston Family Foundation funding awarded to EW, HA, LD, and RV. The funders had no role in study design, collection, or interpretation of data.

## AUTHOR CONTRIBUTIONS

LD, RV, and HA conceived, developed, and coordinated the project. RV, OK, and MS acquired the patient consent. RV, HA, OK, MBJ, SV, MS, FP, KW, DK, SVZ, KK, MG, and LD collected and/or processed stool, mucosal washings and biopsy tissues. RV, SV, HA, NH, MEPM, and MBJ performed and/or analyzed experiments. RV and HA were responsible for the figure preparation and statistical analyses. JW, DG, and EW provided additional supervision and oversight. HA and RV drafted the manuscript. All authors contributed to manuscript edits and approved the final version of the manuscript.

---

**Supplementary Figure 1:**
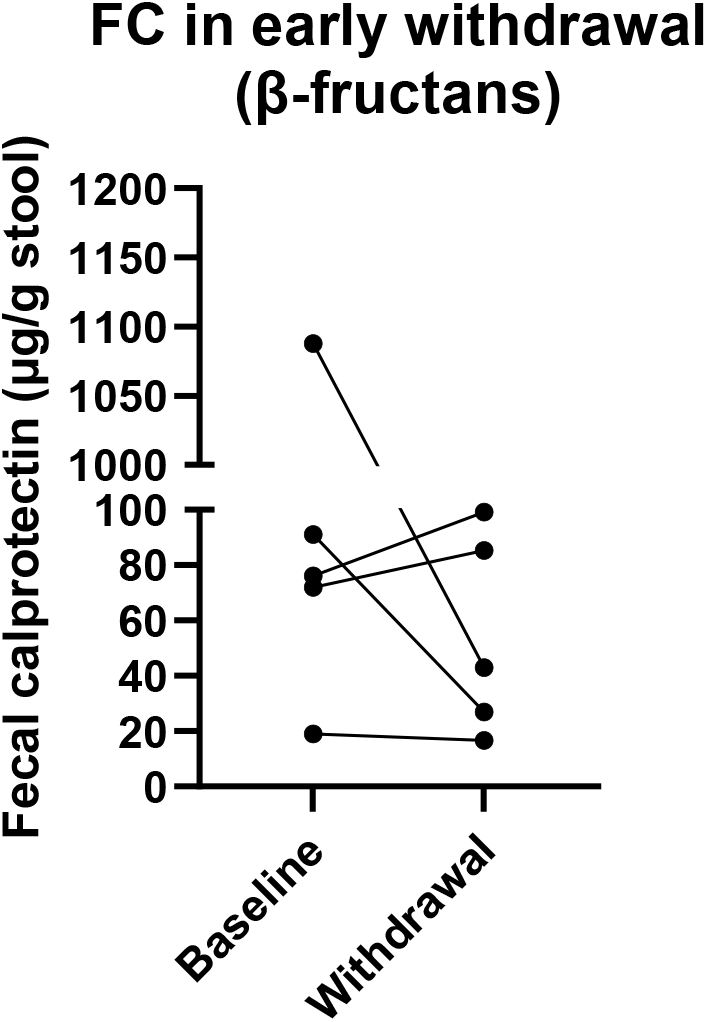
Fecal calprotectin measured at baseline and withdrawal in participants who withdrew from the study early.

